# Incidence of Pulmonary Hypertension in the Echocardiography Referral Population

**DOI:** 10.1101/2024.10.08.24315117

**Authors:** Jonah D. Garry, Suman Kundu, Jeffrey Annis, Chuck Alcorn, Svetlana Eden, Emily Smith, Robert Greevy, Bradley A. Maron, Matthew Freiberg, Evan L. Brittain

**Affiliations:** Division of Cardiovascular Medicine, Vanderbilt University Medical Center, Nashville, TN; Vanderbilt Institute of Clinical and Translational Research, Vanderbilt University Medical Center, Nashville, TN; University of Pittsburgh School of Public Health, Pittsburgh, PA; Department of Biostatistics, Vanderbilt University Medical Center, Nashville, TN; Department of Medicine, University of Maryland School of Medicine, Baltimore, MD and the University of Maryland-Institute for Health Computing, Bethesda, MD; Veterans Affairs Tennessee Valley Healthcare System, Nashville, TN

## Abstract

**Rationale:** Incidence rates for pulmonary hypertension using diagnostic data in patients with cardiopulmonary disease are not known.

**Objectives:** To determine incidence rates of, risk factors for, and mortality hazard associated with pulmonary hypertension among patients referred for transthoracic echocardiography

**Methods:** Retrospective cohort study using data from the Veterans Health Administration (1999- 2020) and Vanderbilt University Medical Center (1994-2020). Pulmonary hypertension was defined as pulmonary artery systolic pressure >35mmHg with prevalent cases excluded. Heart failure and chronic obstructive pulmonary disease were the primary exposures of interest. The primary outcome was incident pulmonary hypertension. Secondarily, we examined mortality rate following incident diagnosis.

**Measurements and Main Results:** We identified 245,067 VA patients (94% male, 20% Black) and 117,526 Vanderbilt patients (46% male, 11% Black) without pulmonary hypertension, of whom 38,882 VA patients and 8,061 Vanderbilt patients developed pulmonary hypertension. Only 18-19% of patients with echo-based pulmonary hypertension also had a diagnostic code. Hazard of pulmonary hypertension was 4-fold higher in patients with heart failure and chronic obstructive pulmonary disease compared to patients without either. Mortality rates increased from pulmonary artery systolic pressure of 35mmHg to 45mmHg then plateaued. Independent risk factors for incident pulmonary hypertension included older age, male sex, black race, and cardiometabolic comorbidities.

**Conclusions:** Pulmonary hypertension incidence rates estimated by diagnostic data are higher than code-based rates. Heart failure and chronic obstructive pulmonary disease strongly associate with incident pulmonary hypertension. Pulmonary artery systolic pressure >45mmHg at diagnosis is associated with high mortality. New pulmonary hypertension on echocardiography is an important prognostic sign.

## Introduction

Pulmonary hypertension (PH) is associated with substantial morbidity and mortality and is most commonly due to heart failure (HF) or chronic obstructive pulmonary disease (COPD) (1, 2). The prevalence of PH in HF is substantial, ranging from 40-75% in HF with reduced left ventricular ejection fraction (HFrEF) and 36-83% in HF with preserved ejection fraction (HFpEF) (3). PH prevalence in COPD is less well described, varying with severity of underlying disease and present in up to 90% of patients with GOLD stage IV disease (4, 5). Despite the notable prevalence of PH and its strong association with increased mortality, it is systemically underrecognized (6, 7). In contrast to virtually all common cardiopulmonary conditions, incident rates (IR) for PH are not well established. Right heart catheterization (RHC) is the gold standard for PH diagnosis, however it is invasive and not routinely performed in all patients with HF or COPD. As a result, estimates of PH incidence in the RHC referral population skew toward sicker, more complex patients. In contrast, transthoracic echocardiogram (TTE) use for estimation of pulmonary pressures has earned widespread acceptance as a non-invasive tool for identifying PH and is routinely used for evaluating symptoms suggestive of PH (8). Moreover, TTE estimated pulmonary artery systolic pressure (PASP) effectively identifies patients at risk of poor outcome, with PASP as low as 33mmHg carrying an increased risk for mortality (9).

Only two prior studies have examined PH IRs (10, 11). The first relied upon on ICD-coding to establish PH diagnosis (10), with findings limited by the unclear accuracy of ICD codes for identifying PH (12, 13). The second used TTE for diagnosis (11), however excluded left-heart disease, the most common cause of PH. To address the gaps in our understanding of PH epidemiology we examined IRs among individuals referred for TTE in a national health care system and a large, sex-balanced tertiary care center. We hypothesized that IRs of PH would be substantially higher than previously reported, particularly among patients with HF and/or COPD, and that incident PH would be associated with increased hazard of mortality.

## Methods

The Institutional Review Boards of the Tennessee Valley Veterans Affairs (VA) Health Care System, West Haven VA Medical Center, and Vanderbilt University Medical Center (Vanderbilt) approved the study. The VA Birth Cohort allows for data analysis through a waiver of consent.

We derived the exploratory cohort from participants in the VA Birth Cohort (14) and evaluated patients with a TTE between September 30, 1999 and October 1, 2020. For the validation cohort we extracted data from Vanderbilt’s de-identified electronic health record. We evaluated patients with a TTE between January 1, 1994 and August 31, 2020. Baseline was defined as the date of the first TTE with a reported PASP value. We excluded patients with PH on baseline TTE. In the VA cohort we excluded patients with baseline TTE after December 31, 2016 to coincide with the end of available Medicare data, which supplies several baseline variables.

### Exposures, Covariates, and Outcomes

We selected HF and COPD as our primary exposures (1). Data regarding baseline clinical characteristics were ascertained using a combination of clinical, laboratory, and or ICD-9/10 code data collected closest to the date of the baseline TTE (up to 180 days after), as previously described (15–21). We used a validated natural language processing tool to extract measurements from TTE reports (22, 23). When absent, we calculated the PASP by simplified Bernoulli equation using the reported tricuspid regurgitant velocity (TRV). We assumed a RAP of 5 mmHg when missing (24, 25). We excluded PASP estimates outside a physiological range. We defined HFpEF as LVEF ≥50% and HFrEF as LVEF <50%, based on LVEF cutoffs frequently used in epidemiologic cohorts and clinical trials (26). Covariate definitions were identical between cohorts, except for smoking and cirrhosis data, which were unavailable in the Vanderbilt cohort.

The primary outcome was incident PH, defined as PASP >35 mmHg, consistent with prior studies (27), near the cutoff suggested by recent guidelines (34.36 mmHg based on TRV of 2.8 m/s and assuming a RAP of 3mmHg) (28), and below 40mmHg (above which PH is consistently liked with adverse outcomes) (29–32). We completed additional sensitivity analyses defining PH by a PASP >40 mmHg. Follow-up time was defined by the time from baseline TTE to follow up TTE with PH, with censoring for death or end of study (September 30th, 2020). In the primary analysis, we conservatively assumed that patients with only one measured PASP value never developed PH. A sensitivity analysis required at least one additional TTE after baseline to define PH incidence. To compare between TTE-based and ICD-based IRs, we calculated PH IRs using ICD-9 and ICD-10 diagnostic codes using the same timeframe as the TTE-based analyses.

The secondary outcome was all-cause mortality after incident PH. In the VA cohort participants were followed from date of incident PH through death or end of study. In the Vanderbilt cohort the mortality analysis was truncated at the end of 2017, after which mortality data is unavailable. Patients who did not develop PH served as a comparator group and were followed from the latest TTE until end of study. Further methodologic details on cohort derivation, exposures, covariates, and outcome analysis are available in the **Online Data Supplement.**

### Statistical Analysis

Baseline characteristics were presented as frequencies and percentages for categorical variables, and median (interquartile range) for continuous variables. We used Poisson regression to estimate IRs per 1000 person-years. Cox proportional hazard regression models were used to estimate the hazard ratios attributable to clinical characteristics. For the mortality outcome, PASP was modeled using restricted cubic splines with three knots in a Cox model. The proportional hazards assumption was tested by the Schoenfeld residuals. We repeated the analysis using time-updated variables of HF, COPD, and eGFR. Kaplan-Meier curves were used to assess the differences in survival among subjects stratified by a 4-level HF and COPD status variable and by PASP category with log-rank testing. All tests with a two-sided p-value <0.05 were considered statistically significant. All missing data was imputed using multiple imputation by chained equations (33). Cox survival models were fitted in each imputed dataset and then combined to obtain pooled hazard ratios and standard errors according to Rubin’s rules (34). All analyses were performed using R software (version 4.0.2; www.r-project.org).

## RESULTS

### VA and Vanderbilt Cohorts

We identified 245,067 VA patients without PH on baseline TTE. Patients were predominantly male (93.7% male; n=15,430 females) with a median age of 59.9 years (IQR 55.1–64.1). Black and Hispanic patients comprised 19.7% and 5.2% of the cohort, respectively. Cardiac, metabolic, and pulmonary comorbidities were common at baseline (**Table 1**), particularly HF (18.2%) and COPD (28.1%). At the end of follow up the prevalence of HF and COPD increased to 36% and 40%, respectively. The median baseline PASP was 28 mmHg (IQR 23–32). A total of 87,007 patients had at least one repeat PASP estimate. The median time between baseline TTE and incident PH was 3.4 years (IQR 1.5–6.4). **Table E1** compares clinical characteristics stratified by the presence of a second TTE after the baseline study and **Table E2** compares clinical characteristics stratified by time between TTEs. There was a higher prevalence of HF (25.5% vs 14.2%) and Diabetes (46.6% vs 38.2%) among individuals with a follow up TTE compared to individuals without. Patients with shorter intervals between TTEs were generally older, with a higher prevalence of HF, COPD, and liver fibrosis (by FIB-4). The proportion of patients who developed incident PH was highest in patients with a follow up TTE within two years (47.6%) compared to longer time intervals (42.3–42.9%).

**Table 1:** Clinical Characteristics of the VA Cohort.

|  | **Overall\***<br>(n=389331) | **Prevalent PH**<br>(n=137939) | **Final analytic cohort†**<br>(n=245067) |
| --- | --- | --- | --- |
| **Age, years (median [IQR])** | 60.1 [55.4, 64.2] | 60.4 [55.8, 64.3] | 59.9 [55.1, 64.1] |
| **Male Sex (%)** | 367653 (94.4) | 132116 (95.8) | 229637 (93.7) |
| **BMI (median [IQR])** | 29.6 [25.8, 34.2] | 29.8 [25.7, 34.8] | 29.5 [25.9, 33.9] |
| **Race/Ethnicity (%)** |  |  |  |
| Other | 12443 (3.2) | 4203 (3.0) | 7986 (3.3) |
| Black | 80364 (20.6) | 31062 (22.5) | 48396 (19.7) |
| Hispanic | 18776 (4.8) | 5525 (4.0) | 12770 (5.2) |
| White | 259145 (66.6) | 89691 (65.0) | 165165 (67.4) |
| **HF (%)** | 102105 (26.2) | 56370 (40.9) | 44637 (18.2) |
| HFrEF (LVEF <50%) | 61122 (15.7) | 35754 (25.9) | 25043 (10.2) |
| HFpEF (LVEF ≥50%) | 40817 (10.5) | 20494 (14.9) | 19594 (8.0) |
| **COPD (%)** | 125722 (32.3) | 55165 (40.0) | 68877 (28.1) |
| **Alcohol abuse (%)** | 103881 (26.7) | 39616 (28.7) | 62668 (25.6) |
| **Cocaine abuse (%)** | 32541 (8.4) | 12267 (8.9) | 19880 (8.1) |
| **Scleroderma (%)** | 11570 (3.0) | 4436 (3.2) | 6942 (2.8) |
| **Smoking (%)** |  |  |  |
| Current | 147385 (37.9) | 54490 (39.5) | 77130 (31.5) |
| Former | 117090 (30.1) | 42116 (30.5) | 57897 (23.6) |
| Never | 113694 (29.2) | 36911 (26.8) | 56503 (23.1) |
| **Hypertension (%)** |  |  |  |
| None | 10817 (2.8) | 2797 (2.0) | 7757 (3.2) |
| Controlled | 287625 (73.9) | 99357 (72.0) | 183639 (74.9) |
| Uncontrolled | 77558 (19.9) | 30803 (22.3) | 45609 (18.6) |
| **Diabetes Mellitus (%)** | 170748 (43.9) | 67588 (49.0) | 100935 (41.2) |
| **Hepatitis C Infection (%)** | 47028 (12.1) | 17802 (12.9) | 28558 (11.7) |
| **FIB-4 >3.25** | 32359 (8.3) | 14125 (10.2) | 13980 (5.7) |
| **eGFR (median [IQR])** | 82.0 [66.0, 98.0] | 80.0 [61.0, 97.0] | 85.0 [71.0, 101.0] |
| **Hgb (median [IQR])** | 14.0 [12.5, 15.1] | 13.5 [11.7, 14.8] | 14.20 [12.9, 15.2] |

BMI = Body Mass Index
HF = Heart Failure
eGFR = estimated Glomerular Filtration Rate
Hgb = Hemoglobin
\*(All with baseline PASP value before 12/2016)
†Missing values in the final analytic cohort n (%): Race: 10858 [4.4%]; Smoking status: 53714 [21.9%];
Hypertension: 8201 [3.3%]; eGFR : 2394 [1%]; BMI: 3332 [1.4%]; FIB-4: 8122 [3.3%]; Hemoglobin 3522 [1.4%]

We identified 77,548 Vanderbilt patients without PH on baseline TTE. Similar to the VA cohort, HF (21.4%) and COPD (12.3%) were common in the Vanderbilt cohort, as were other comorbidities (**Table 2**).

**Table 2:** Clinical Characteristics of the Vanderbilt Cohort.

|  | **Overall**<br>(n=117526) | **Prevalent PH**<br>(n=36586) | **Final analytic cohort\***<br>(n=77548) |
| --- | --- | --- | --- |
| **Age, years (median [IQR])** | 61.8 [48.5, 72.0] | 66.9 [55.6, 76.4] | 59.6 [46.0, 69.9] |
| **Male Sex (%)** | 54847 (46.7) | 17488 (47.8) | 35775 (46.1) |
| **BMI (median [IQR])** | 28.2 [24.3, 33.1] | 28.4 [24.4, 33.8] | 28.1 [24.2, 32.9] |
| **Race/Ethnicity (%)** |  |  |  |
| Other | 8262 (7.0) | 2503 (6.8) | 4783 (6.2) |
| Black | 14178 (12.1) | 5251 (14.4) | 8634 (11.1) |
| Hispanic | 1721 (1.5) | 403 (1.1) | 1251 (1.6) |
| White | 93365 (79.4) | 28429 (77.7) | 62880 (81.1) |
| **HF (%)** | 34810 (29.6) | 17907 (48.9) | 16603 (21.4) |
| HF<sub>r</sub>EF (LVEF <50%) | 14389 (12.2) | 8050 (22.0) | 6235 (8.0) |
| HF<sub>p</sub>EF (LVEF ≥50%) | 19473 (16.6) | 9300 (25.4) | 9984 (12.9) |
| **COPD (%)** | 18344 (15.6) | 8624 (23.6) | 9549 (12.3) |
| **Alcohol abuse (%)** | 5280 (4.5) | 1809 (4.9) | 3399 (4.4) |
| **Cocaine abuse (%)** | 1212 (1.0) | 407 (1.1) | 793 (1.0) |
| **Scleroderma (%)** | 5211 (4.4) | 1852 (5.1) | 3301 (4.3) |
| **Smoking (%)** |  |  |  |
| Current Smoker | 7654 (6.5) | 2171 (5.9) | 5219 (6.7) |
| Former Smoker | 17773 (15.1) | 5644 (15.4) | 11759 (15.2) |
| Never Smoker | 34755 (29.6) | 8546 (23.4) | 25163 (32.4) |
| **Hypertension (%)** |  |  |  |
| None | 9949 (8.5) | 1020 (2.8) | 7926 (10.2) |
| Controlled | 75173 (64.0) | 23460 (64.1) | 50336 (64.9) |
| Uncontrolled | 21677 (18.4) | 7780 (21.3) | 13418 (17.3) |
| **Diabetes Mellitus (%)** | 16050 (13.7) | 6197 (16.9) | 9798 (12.6) |
| **Hepatitis C Infection (%)** | 3365 (2.9) | 1106 (3.0) | 2234 (2.9) |
| **FIB-4 >3.25** | 12817 (10.9) | 5758 (15.7) | 6918 (8.9) |
| **eGFR (median [IQR])** | 77.0 [57.0, 97.0] | 67.0 [45.0, 88.5] | 81.0 [63.0, 100.0] |
| **Hgb (median [IQR])** | 12.50 [10.60, 13.90] | 11.60 [9.80, 13.30] | 12.80 [11.10, 14.10] |

BMI = Body Mass Index
HF = Heart Failure
eGFR = estimated Glomerular Filtration Rate
Hgb = Hemoglobin
**\*Missing values in the final analytic cohort n (%):** Gender: 6 [<0.01%]; Smoking: 35407 [45.7%]; Hypertension: 5868 [7.6%]; FIB-4: 28617 [36.9%]; eGFR: 12673 [16.3%]; BMI: 9658 [12.5%]; LVEF: 384 [0.5%]; Hemoglobin: 14779 [19.1%]

### PH Incidence Rates

Incident PH developed in 38,882 (15.9%) VA patients over a median follow up of 6.6 years (IQR 4.1–10.2) and 8,061 (10.4%) Vanderbilt patients over a median follow up of 2.7 years (IQR 0.8–6.2). The median change in PASP in the VA cohort was 14.0 mmHg (IQR 8.4– 21.7) and in the Vanderbilt cohort was 13.1 mmHg (IQR 8.3–19.9). A total of 39,741 VA patients (16.2%) and 9,845 Vanderbilt patients (12.7%) had an increase of 5 mmHg or more between TTEs. PASP was 32-34 mmHg on baseline TTE in 13.1% of the VA cohort and 8.2% of the Vanderbilt cohort. Patients with 32-34mmHg on their baseline TTE comprised 17% of incident PH cases in the VA cohort and 12.6% of cases in the Vanderbilt cohort.

The IR of PH in the VA cohort was 22.0 per 1000 person-years. HF, COPD, or both were present at baseline in 51% of patients who developed incident PH. IRs were lower for COPD alone than HF alone and lowest among those with neither HF nor COPD (**Table 3, Figure E1**). The highest IRs were observed among those with both HF and COPD (60.3 vs 15.7 per 1000 person-years). Among patients with HF at baseline, PH IRs were higher in those with HFrEF compared with HFpEF (62.7 vs 43.9 per 1000 person-years). We observed similar but consistently higher PH IRs in the Vanderbilt validation cohort compared to the VA cohort (**Table 3**). Among the 76,007 VA patients and 21,653 Vanderbilt patients with at least one subsequent PASP measurement IRs were 2-3 fold higher (**Table E3**).

**Table 3:** PH IRs in the VA and Vanderbilt Cohorts, stratified by HF/COPD Status and within HF by LVEF.

|  | VA PH IRs |  |  | Vanderbilt PH IRs |  |  |
| --- | --- | --- | --- | --- | --- | --- |
|  | N | Incident PH | Rate/1000 PY | N | Incident PH | Rate/1000 PY |
| **Total** | 245067 | 38882 | 22.0 (21.8, 22.3) | 77548 | 8061 | 25.5 (24.9, 26.0) |
| **HF-/COPD-** | 152456 | 19111 | 15.7 (15.5, 16.0) | 55258 | 4199 | 17.2 (16.7, 17.7) |
| **HF-/COPD+** | 47974 | 7128 | 22.6 (22.1, 23.1) | 5687 | 555 | 31.4 (28.9, 34.1) |
| **HF+/COPD-** | 23734 | 6856 | 49.8 (48.6, 51.0) | 12741 | 2524 | 56.4 (54.2, 58.6) |
| **HF+/COPD+** | 20903 | 5787 | 60.3 (58.8, 61.9) | 3862 | 783 | 79.8 (74.3, 85.5) |
| **HF+/LVEF <50%** | 25043 | 7966 | 62.7 (61.3, 64.1) | 6235 | 1478 | 78.2 (74.3 - 82.2) |
| **HF+/LVEF ≥50%** | 18594 | 4677 | 43.9 (42.7, 45.2) | 9984 | 1751 | 51.0 (48.6 - 53.4) |

HF = Heart Failure
COPD = Chronic Obstructive Pulmonary Disease
LVEF = Left Ventricular Ejection Fraction

PH IRs did not vary over time in the VA cohort (**Table E4**) when stratified by five-year intervals beginning in 2000. PH IRs were approximately 30-40% lower when PH was defined by PASP>40mmHg (**Table E5**) in both cohorts. Among patients with TTE-based incident PH, only 19% of VA cohort and 18% of the Vanderbilt cohort also had an ICD-code for PH (**Table E6**). When defined by ICD-coding, the PH IRs were lower in both the VA (7.5 vs 22.0 per 1000 person-years) and Vanderbilt cohorts (5.9 vs 25.5 per 1000 person-years) (**Table E7**).

### Clinical Characteristic Associations

Within the VA cohort, there was an increased hazard of incident PH with COPD alone (HR 1.30; 95% CI 1.26-1.33), HF alone (HR 2.38; 95% CI 2.31-2.44) and concurrent HF and COPD (HR 2.54, 95% CI 2.46-2.62) in comparison to individuals without HF or COPD. The hazard for incident PH associated with HF and COPD increased substantially after time-updating these comorbidities **(Table 4**). Among individuals with HF, HFrEF was associated with greater risk of incident PH compared to HFpEF (HR 1.44; 95% CI 1.39-1.49). Hazard of PH increased with age, hypertension, diabetes, and atrial fibrillation. We found higher risk of PH among males (HR 1.40; 95% CI 1.33-1.47, p<0.001) and non-Hispanic Black patients (HR 1.04; 95% CI 1.01- 1.07, p<0.001). The covariates associated with incident PH in the Vanderbilt cohort were similar to the VA cohort (**Table 4**). Excluding smoking history and FIB-4 covariates (absent in the VUMC cohort) from the VA cohort did not change the significance of any covariate associations. The factors associated with incident PH were unchanged on sensitivity analysis defining PH as PASP>40 mmHg (**Table E8**).

**Table 4:** Predictors of Incident PH in the VA and Vanderbilt Cohorts.

| Characteristic | VA Cohort |  | Vanderbilt Cohort |  |
| --- | --- | --- | --- | --- |
|  | Hazard Ratio [95% CI] |  | Hazard Ratio [95% CI] |  |
|  | Baseline | Time updated * | Baseline | Time updated * |
| **HF-/COPD-** | 1 | 1 | 1 | 1 |
| **HF-/COPD+** | 1.30 (1.26-1.33) | 1.52 (1.47-1.57) | 1.38 (1.26-1.51) | 1.57 (1.43-1.73) |
| **HF+/COPD-** | 2.38 (2.31-2.44) | 3.25 (3.14-3.35) | 2.27 (2.15-2.39) | 2.93 (2.77-3.09) |
| **HF+/COPD+** | 2.54 (2.46-2.62) | 4.07 (3.95-4.20) | 2.60 (2.40-2.82) | 3.63 (3.37-3.91) |
| **Age, years (per SD, 6.4)** | 1.12 (1.10-1.13) | 1.07 (1.06-1.08) | 1.29 (1.26-1.33) | 1.18 (1.15-1.21) |
| **Male Sex** | 1.40 (1.33-1.47) | 1.32 (1.25-1.39) | 1.14 (1.09-1.20) | 1.11 (1.06-1.17) |
| **BMI, kg/m<sup>2</sup> (per SD, 6.4)** | 1.01 (1.00-1.02) | 0.98 (0.97-0.99) | 0.99 (0.94-1.05) | 0.99 (0.94-1.05) |
| **Race/Ethnicity (vs. Non-Hispanic White)** |  |  |  |  |
| Other | 0.95 (0.90-1.01) | 0.95 (0.90-1.00) | 0.76 (0.67-0.88) | 0.73 (0.63-0.85) |
| Non-Hispanic Black | 1.04 (1.01-1.07) | 1.06 (1.03-1.09) | 1.18 (1.10-1.26) | 1.15 (1.08-1.24) |
| Hispanic | 0.92 (0.88-0.97) | 0.95 (0.90-1.00) | 0.82 (0.66-1.02) | 0.82 (0.65-1.03) |
| **Hypertension (vs. No Hypertension)** |  |  |  |  |
| *Controlled Hypertension* | 2.33 (2.07-2.62) | 2.04 (1.79-2.32) | 1.49 (1.31-1.70) | 1.32 (1.15-1.50) |
| *Uncontrolled Hypertension* | 2.72 (2.41-3.06) | 2.25 (1.98-2.56) | 1.60 (1.38-1.85) | 1.38 (1.20-1.60) |
| **Diabetes Mellitus** | 1.31 (1.28-1.34) | 1.17 (1.14-1.19) | 1.30 (1.23-1.38) | 1.13 (1.07-1.20) |
| **Veterans with HIV** | 1.12 (1.03-1.23) | 1.04 (0.94-1.14) | 0.98 (0.82-1.17) | 0.91 (0.76-1.10) |
| **Hepatitis C Infection** | 1.24 (1.20-1.28) | 1.21 (1.17-1.25) | 1.36 (1.19-1.55) | 1.30 (1.13-1.49) |
| **Smoking Status (vs. Never)** |  |  | - | - |
| *Current* | 1.17 (1.13-1.20) | 1.08 (1.05-1.12) | - | - |
| *Former* | 1.09 (1.06-1.12) | 1.05 (1.02-1.09) | - | - |
| **Liver Fibrosis (FIB-4 >3.25 vs. ≤325)** | 1.34 (1.28-1.40) | 1.36 (1.30-1.43) | - | - |
| **eGFR, mL/min/1.73m<sup>2</sup> (per SD, 2.9)** | 0.89 (0.88-0.90) | 0.75 (0.73-0.76) | 0.82 (0.80-0.85) | 0.67 (0.65-0.70) |
| **Alcohol abuse** | 1.03 (1.00-1.05) | 1.01 (0.98-1.04) | 0.97 (0.86-1.10) | 0.95 (0.84-1.08) |
| **Scleroderma** | 1.18 (1.12-1.25) | 1.15 (1.09-1.22) | 1.25 (1.14-1.38) | 1.23 (1.11-1.36) |
| **Cocaine abuse** | 1.01 (0.96-1.05) | 0.98 (0.93-1.02) | 1.30 (1.05-1.60) | 1.26 (1.01-1.57) |
| **Atrial fibrillation** | 1.64 (1.60-1.69) | 1.51 (1.47-1.55) | 1.59 (1.51-1.67) | 1.50 (1.42-1.58) |
| **Hgb, mg/dL (per SD, 1.9)** | 0.84 (0.83-0.85) | 0.89 (0.88-0.90) | 0.83 (0.81-0.85) | 0.86 (0.83-0.88) |

\*HF, COPD, and eGFR and are time updated. Other variables measured solely at baseline
HF = Heart Failure
COPD = Chronic Obstructive Pulmonary Disease
BMI = Body Mass Index
eGFR = estimated Glomerular Filtration Rate
Hgb = Hemoglobin

### Mortality Risk

In the VA cohort 16,238 patients died over a median follow up of 3.4 years (IQR 1.4-6.4). After incident PH, patients with concurrent HF and COPD had an approximately 2.5-fold higher mortality rate compared to those with neither HF nor COPD while mortality rate was higher in patients with HF alone than COPD alone. The increased hazard of mortality attributable to HF, COPD, or both in comparison to patients with neither was proportionally similar in the Vanderbilt and VA cohorts **(Figure 1, Table E9)**. Mortality rates based on incident PASP demonstrate a large increase in risk between 35-45mmHg and 45-55mmHg, above which rates increase only modestly (**Table E10**). This finding was robust regardless of HF and COPD status **(Figure E2)** or time between baseline TTE and incident PH TTE (**Figure E3**). When examined continuously, we found hazard of mortality linearly increased from 35mmHg up to 45mmHg, after which the hazard plateaued in both the VA and Vanderbilt Cohorts (**Figure 2, Figure E4)**.

**Figure 1:**
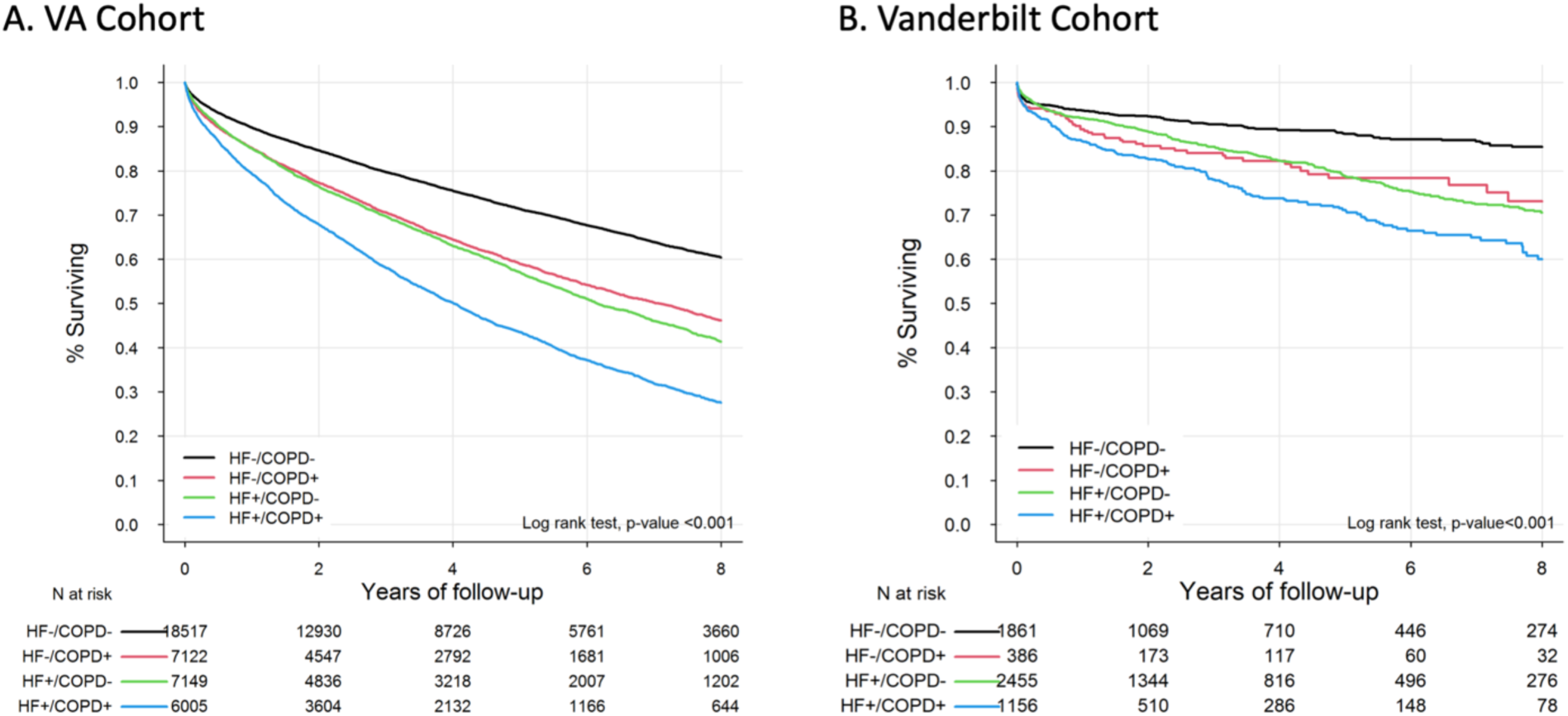
Survival following Incident PH Diagnosis by Comorbid HF/COPD Status. Displayed are the Kaplan-Meier time-to-event curves for survival in both the VA cohort (A) and the Vanderbilt cohorts (B), with stratification by HF and COPD status. Concurrent HF and COPD are associated with the highest unadjusted rate of mortality, followed by the presence of HF or COPD alone, with the lowest rate of mortality in individuals without HF or COPD. Results are similar in both cohorts.

**Figure 2:**
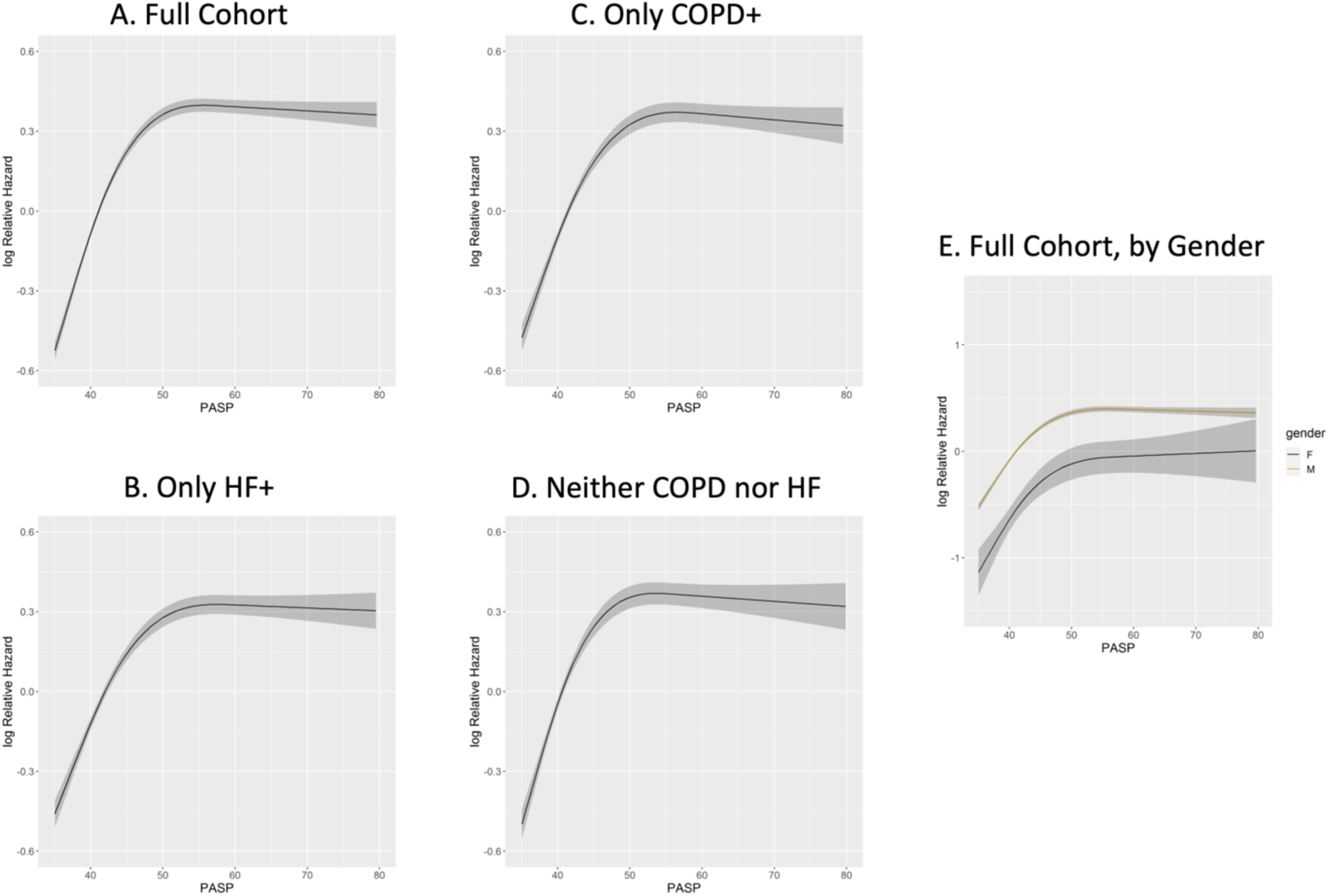
Mortality hazard by PASP at incident PH diagnosis (VA Cohort) We evaluated the relationship between PASP at the time of echocardiographic incident PH diagnosis and hazard of mortality by restricted cubic splines with up to 3 knots in the VA cohort. We assessed this relationship in the full cohort (A), patients with HF and no COPD (B), patients with COPD and no HF (C), and patients with neither COPD nor HF (D). In all groups, the relative hazard of mortality increases linearly as PASP increases from 35 mmHg up to approximately 45- 50mmHg, above which hazard of mortality remains stable. We then stratified the full cohort by gender (E) and found that mortality risk plateaus for men at approximately 45-50mmHg, but continues to gradually increase for women.

## Discussion

Our study substantially expands upon prior PH incidence and prevalence studies, using raw diagnostic information from clinical TTEs and assessing comorbidity specific rates in two large racially and ethnically diverse cohorts. The major findings are: 1) The IRs of PH in the TTE referral population are substantially higher than previously appreciated by code-based definitions. 2) Both HF and COPD increase the hazard of incident PH (193%-225% and 52-57% increased risk, respectively) with concurrent diagnosis of HF and COPD associated with additive risk (263%-307%). 3) The predominant risk factors for incident PH independent of COPD and HF include age, male sex, black race, and cardiometabolic comorbidities. 4) Incident PH is associated with increased hazard of mortality. This hazard increases with comorbid COPD or HF as well as higher PASP to approximately 45mmHg. This work provides important new evidence for PH surveillance among at-risk individuals and identifies hemodynamic thresholds associated with highest risk in patients with newly diagnosed PH.

The only prior study to describe PH IRs across WSPH groups estimated the PH incidence to be 19.8-24.1 per 100,000 patient-years in a population-wide regional Canadian healthcare database relying upon ICD-codes (10). We identified a PH IR estimate approximately 30-fold higher among patients referred for TTE. In contrast to prior findings, our data suggest that the IR of PH is stable over time, aligning with epidemiologic evidence that the IRs of HF and COPD are stable or decreasing (35–37). Code-based incident PH was diagnosed in 18-19% of TTE- diagnosed cases with IRs one-fourth to one-half as high as TTE-based rates. This is likely due to clinical under-recognition, as previously noted (38). Conversely, approximately 50% of cases identified by ICD-coding were not identified by TTE, which may be due to unrecognized cases, but more likely due inappropriate coding or suspicion for PH that is not subsequently confirmed (38). The magnitude of difference in IRs between the studies may be explained by enrichment for cardiopulmonary comorbidities (among others) in the TTE referral population compared with the general population. A TTE referral cohort may have lower generalizability to the overall population but is highly relevant to patients seeking care and their providers. This may be particularly impactful in the HF and COPD populations in which the minority of patients undergo RHC (39), a practice consistent with current PH guidelines when there is little uncertainty of the underlying etiology (40).

In patients with at least one follow-up TTE after baseline, we found lower PH IRs (65.9- 69.1 per 1000 person-years) in comparison to one prior study examining TTE-based PH IRs (92.3 per 1000 person-years) (11). Variation between the estimates may be explained by differing definitions of PH (PASP≥30mmHg vs PASP>35mmHg) and exclusion of patients with left heart disease. Our study reinforces and expands upon Stewart et al (11) through larger, U.S.- based, racially and ethnically diverse cohorts inclusive of left heart disease, and clarifies the contribution of comorbidities to hazard of Incident PH.

Prior TTE-based studies of prevalent PH within population-based initiatives have noted increased prevalence of PH among older adults and patients with comorbidities (27, 41, 42). We found that HF was the strongest single factor associated with incident PH, with HFrEF more strongly associated than HFpEF. Within the HF population LV dysfunction may associate with higher left-sided filling pressures and thereby degree of pulmonary venous congestion (43), however information on LV filling pressure was not available in our study.

COPD was strongly associated with incident PH, correlating with prior evidence of PH prevalence in COPD estimated at 40% in RHC referral cohorts, although these may be biased toward more severe cases (44, 45). The relationship between severity of COPD and hazard of incident PH remains an outstanding question. In patients with concurrent HF and COPD the hazard of incident PH was additive, suggesting distinct pathophysiologic processes contributing to PH.

Age, male sex, and Black race were independently associated with increased hazard of incident PH. Pulmonary artery pressure increases with age and is hypothesized to occur through increased blood vessel stiffness and impaired LV diastolic function (43, 46–48). Several studies have suggested that the prevalence of PH is higher in women than men across WHO groups (39, 49), however findings from ARIC suggest no difference in PASP change over time between sexes (43) and we found that male sex was associated with increased hazard of incident PH. Based upon these discordant findings the population-level association between sex and PH incidence remains unclear. We found an increased hazard of incident PH among patients who self-identified as non-Hispanic Black race compared to White race, aligning with prior analyses in prevalent PH (50, 51) but conflicting with ARIC findings of no differences in PASP change over time between Black and White participants (43). Overall, the differing findings from our cohorts and the ARIC cohort may be explained by substantial differences in patient characteristics. Cardiometabolic risk factors including atrial fibrillation, hypertension, and diabetes were associated with incident PH, building upon prior associations noted in prevalent PH cohorts (52, 53).

We found a strong association between incident PH and increased hazard of all-cause mortality, building upon prior studies (27, 42, 54) We provide new granularity by modeling PASP continuously, rather than categorizing into ranges. We found that mortality hazard increases linearly with pulmonary pressure until ∼45 mmHg, above which the hazard plateaus.

Further, we found the relationship between RVSP and mortality above 50 mmHg may vary by gender with a more significant plateau in mortality risk among men than women. This may reflect gender differences in right ventricular compensation, differing etiologies of PH between the genders, or other unaccounted for co-associating factors. These findings are consistent with prior studies in prevalent PH, with ∼45-50 mmHg possibly representing a threshold for RV dysfunction (55, 56). We contend that incident PH should be recognized as a powerful prognostic indicator, especially when PASP is >45 mmHg. PH is likely associated with mortality directly through right ventricular dysfunction and indirectly as a marker of comorbid disease severity.

### Strengths

The cohorts examined in this study are the largest to-date describing risk factors and risk of mortality in incident PH. The use of TTE to identify PH is more clinically practical than other diagnostic modalities and is reasonably accurate. Our findings were consistent in two distinct cohorts that were similar in their demographics and prevalence of comorbidities with racial and ethnic representation reflective of the U.S. population. We performed multiple sensitivity analyses to bolster the validity of our findings, accounting for time-updated comorbid diagnoses and the effect of time between TTEs on risk of mortality, reporting PH IR over a 20-year period, defining PH with multiple cutoffs, and restricting the cohort to individuals with a second TTE.

### Limitations

This is an observational study and our findings do not imply causality, but rather highlight the importance of future mechanistic work. Referral cohorts are inherently different than the general population, therefore all results must be interpreted within the context of the populations studied. Although we adjusted for a wide range of confounders we cannot rule out the possibility of residual confounding from variables that could not be measured. Reflective of a real-world clinical population the time between studies was not standardized, and it is possible that PH was systematically identified late. Indication for TTE also varied and could not be ascertained, which also may introduce bias. Loss to follow up could not be estimated, therefore we assumed patients without a second TTE to have not developed PH, which provides a conservative estimate. In contrast, our sensitivity analysis requiring at least one follow-up TTE after baseline likely overestimates IRs compared with the overall referral population. Outside of protocolized enrollment, the VA as a closed healthcare system offers the best opportunity to capture all follow up compared to other US healthcare systems. TTE is not the gold standard for diagnosing PH and cannot differentiate between WHO groups of PH (57). The minimal detectable difference for PASP is not well established and has only been examined in a small cohort of systemic sclerosis (58), which may limit the interpretability of our findings for individual patients with borderline PASP values, however these patients represent a small proportion of incident PH cases. Our VA cohort age range is limited to patients born between 1945-1965 with a TTE 1999-2020 (minimum age 34 years, maximum age 75). This may introduce bias by excluding younger patients with WSPH Group 1 PH and older patients at higher risk of incident PH. This limitation was not present in our Vanderbilt cohort, which yielded similar findings.

## Conclusions

Incident PH is markedly more common among the TTE referral population than previously recognized. Comorbid HF and COPD are strongly associated with incident PH, along with demographic factors and cardiometabolic comorbidities. Individuals with incident PH are at increased risk of mortality and new PH on TTE should be recognized as a significant prognostic sign. There is a substantial population of individuals at risk for PH in need of preventative strategies and therapeutics.

## Data Availability

All data produced in the present study are available upon reasonable request to the authors.

## Non-standard Abbreviations and Acronyms

BMI: Body Mass Index
COPD: Chronic Obstructive Pulmonary Disease
eGFR: estimated glomerular filtration rate
HF: Heart Failure
HFrEF: Heart Failure with reduced ejection fraction
HFpEF: Heart Failure with preserved ejection fraction
IR: Incident Rate
LV: Left ventricular
LVEF: Left Ventricular ejection fraction
PASP: Pulmonary Artery Systolic Pressure
PH: Pulmonary Hypertension
RAP: Right atrial pressure
RHC: Right Heart Catheterization
SSDI: Social Security Death Index
TTE: Transthoracic Echocardiogram
VA: Veterans Affairs

## Acknowledgments

This work uses data provided by patients and collected by the VA as part of their care and support.

## VA Disclaimer

The views and opinions expressed in this manuscript are those of the authors and do not necessarily represent those of the Department of Veterans Affairs or the United States government

## Notes

**Support:** This work was supported by the National Heart Lung and Blood Institute: T32 HL 087738 (Garry), R01 HL 163960 (Brittain), R01 HL 146588 (Brittain, Freiberg), R01 HL 155278 (Brittain); National Institute of Diabetes and Digestive and Kidney Diseases: R01 DK 124845 (Brittain); and US Food and Drug Administration R01 FD 007627 (Brittain). The Veterans Affairs Cohort Study is supported by P01 AA 029545 and U24 AA 020794.

### Competing Interest Statement

The authors have declared no competing interest.

### Funding Statement

This study was funded by the National Heart Lung and Blood Institute: T32 HL 087738 (Garry), R01 HL 163960 (Brittain), R01 HL 146588 (Brittain, Freiberg), R01 HL 155278 (Brittain); National Institute of Diabetes and Digestive and Kidney Diseases: R01 DK 124845 (Brittain); and US Food and Drug Administration R01 FD 007627 (Brittain). The Veterans Affairs Cohort Study is supported by P01 AA 029545 and U24 AA 020794.

### Author Declarations

The Institutional Review Boards of the Tennessee Valley Veterans Affairs (VA) Health Care System, West Haven VA Medical Center, and Vanderbilt University Medical Center gave ethical approval for this work.

## References

1. Hoeper MM, Humbert M, Souza R, Idrees M, Kawut SM, Sliwa-Hahnle K, et al. A global view of pulmonary hypertension. Lancet Respir Med 2016;

2. Strange G, Stewart S, Celermajer DS, Prior D, Scalia GM, Marwick TH, et al. Threshold of Pulmonary Hypertension Associated With Increased Mortality. J Am Coll Cardiol 2019;73:2660– 2672.

3. Rosenkranz S, Gibbs JSR, Wachter R, De Marco T, Vonk-Noordegraaf A, Vachiéry JL. Left ventricular heart failure and pulmonary hypertension. Eur Heart J 2016;37:942–954.

4. Seeger W, Adir Y, Barberà JA, Champion H, Coghlan JG, Cottin V, et al. Pulmonary hypertension in chronic lung diseases. J Am Coll Cardiol 2013;62:.

5. Nathan SD, Barbera JA, Gaine SP, Harari S, Martinez FJ, Olschewski H, et al. Pulmonary hypertension in chronic lung disease and hypoxia. Eur Respir J 2019;53:.

6. Jankowich M, Maron BA, Choudhary G. Mildly elevated pulmonary artery systolic pressure on echocardiography: bridging the gap in current guidelines. Lancet Respir Med 2021;9:1185–1191.

7. Maron BA, Choudhary G, Khan UA, Jankowich MD, McChesney H, Ferrazzani SJ, et al. Clinical profile and underdiagnosis of pulmonary hypertension in US veteran patients. Circ Heart Fail 2013;6:906–912.

8. Humbert M, Kovacs G, Hoeper MM, Badagliacca R, Berger RMF, Brida M, et al. 2022 ESC/ERS Guidelines for the diagnosis and treatment of pulmonary hypertension. Eur Respir J 2023;61:.

9. Huston JH, Maron BA, French J, Huang S, Thayer T, Farber-Eger EH, et al. Association of Mild Echocardiographic Pulmonary Hypertension With Mortality and Right Ventricular Function. JAMA Cardiol 2019;4:1112–1121.

10. Thiwanka Wijeratne D, Lajkosz K, Brogly SB, Diane Lougheed M, Jiang L, Housin A, et al. Increasing Incidence and Prevalence of World Health Organization Groups 1 to 4 Pulmonary Hypertension: A Population-Based Cohort Study in Ontario, Canada. Circ Cardiovasc Qual Outcomes 2018;11:.

11. Stewart S, Chan Y-K, Playford D, Harris S, Strange GA. Incident pulmonary hypertension in 13 488 cases investigated with repeat echocardiography: a clinical cohort study. ERJ Open Res 2023;9:.

12. Link J, Glazer C, Torres F, Chin K. International Classification of Diseases Coding Changes Lead to Profound Declines in Reported Idiopathic Pulmonary Arterial Hypertension Mortality and Hospitalizations: Implications for Database Studies. Chest 2011;139:497–504.

13. Mathai SC, Mathew S. Breathing (and Coding?) a Bit Easier: Changes to International Classification of Disease Coding for Pulmonary Hypertension. Chest 2018;154:207–218.

14. Sarkar S, Esserman DA, Skanderson M, Levin FL, Justice AC, Lim JK. Disparities in hepatitis C testing in U.S. veterans born 1945-1965. J Hepatol 2016;65:259–265.

15. McGinnis KA, Brandt CA, Skanderson M, Justice AC, Shahrir S, Butt AA, et al. Validating smoking data from the Veteran’s Affairs Health Factors dataset, an electronic data source. Nicotine Tob Res 2011;13:1233–1239.

16. Butt AA, McGinnis K, Rodriguez-Barradas MC, Crystal S, Simberkoff M, Goetz MB, et al. HIV infection and the risk of diabetes mellitus. AIDS 2009;23:1227–1234.

17. McGinnis KA, Justice AC, Bailin S, Wellons M, Freiberg M, Koethe JR. High concordance between chart review adjudication and electronic medical record data to identify prevalent and incident diabetes mellitus among persons with and without HIV. Pharmacoepidemiol Drug Saf 2020;29:1432–1439.

18. Chobanian A V., Bakris GL, Black HR, Cushman WC, Green LA, Izzo JL, et al. Seventh report of the Joint National Committee on Prevention, Detection, Evaluation, and Treatment of High Blood Pressure. Hypertension 2003;42:1206–1252.

19. Fultz SL, Skanderson M, Mole LA, Gandhi N, Bryant K, Crystal S, et al. Development and verification of a “virtual” cohort using the National VA Health Information System. Med Care 2006;44:.

20. Maron BA, Hess E, Maddox TM, Opotowsky AR, Tedford RJ, Lahm T, et al. Association of Borderline Pulmonary Hypertension With Mortality and Hospitalization in a Large Patient Cohort: Insights From the Veterans Affairs Clinical Assessment, Reporting, and Tracking Program. Circulation 2016;133:1240–1248.

21. Crothers K, Huang L, Goulet JL, Goetz MB, Brown ST, Rodriguez-Barradas MC, et al. HIV infection and risk for incident pulmonary diseases in the combination antiretroviral therapy era. Am J Respir Crit Care Med 2011;183:388–395.

22. Patterson O V., Freiberg MS, Skanderson M, Fodeh JS, Brandt CA, DuVall SL. Unlocking echocardiogram measurements for heart disease research through natural language processing. BMC Cardiovasc Disord 2017;17:.

23. Wells QS, Farber-Eger E, Crawford DC. Extraction of echocardiographic data from the electronic medical record is a rapid and efficient method for study of cardiac structure and function. J Clin Bioinforma 2014;4:.

24. Lam CSP, Roger VL, Rodeheffer RJ, Borlaug BA, Enders FT, Redfield MM. Pulmonary hypertension in heart failure with preserved ejection fraction: a community-based study. J Am Coll Cardiol 2009;53:1119–1126.

25. Choudhary G, Jankowich M, Wu WC. Elevated pulmonary artery systolic pressure predicts heart failure admissions in African Americans jackson heart study. Circ Heart Fail 2014;7:558–564.

26. Heidenreich PA, Bozkurt B, Aguilar D, Allen LA, Byun JJ, Colvin MM, et al. 2022 AHA/ACC/HFSA Guideline for the Management of Heart Failure: A Report of the American College of Cardiology/American Heart Association Joint Committee on Clinical Practice Guidelines. Circulation 2022;

27. Lam CSP, Borlaug BA, Kane GC, Enders FT, Rodeheffer RJ, Redfield MM. Age-associated increases in pulmonary artery systolic pressure in the general population. Circulation 2009;119:2663–2670.

28. Humbert M, Kovacs G, Hoeper MM, Badagliacca R, Berger RMF, Brida M, et al. 2022 ESC/ERS Guidelines for the diagnosis and treatment of pulmonary hypertension. Eur Heart J 2022;43:3618–3731.

29. Kolte D, Lakshmanan S, Jankowich MD, Brittain EL, Maron BA, Choudhary G. Mild pulmonary hypertension is associated with increased mortality: A systematic review and meta-analysis. J Am Heart Assoc 2018;7:.

30. Douschan P, Kovacs G, Avian A, Foris V, Gruber F, Olschewski A, et al. Mild elevation of pulmonary arterial pressure as a predictor of mortality. Am J Respir Crit Care Med 2018;197:509– 516.

31. Assad TR, Maron BA, Robbins IM, Xu M, Huang S, Harrell FE, et al. Prognostic Effect and Longitudinal Hemodynamic Assessment of Borderline Pulmonary Hypertension. JAMA Cardiol 2017;2:1361–1368.

32. Simonneau G, Montani D, Celermajer DS, Denton CP, Gatzoulis MA, Krowka M, et al. Haemodynamic definitions and updated clinical classification of pulmonary hypertension. European Respiratory Journal 2019;53:.

33. van Buuren S, Groothuis-Oudshoorn K. mice: Multivariate Imputation by Chained Equations in R. J Stat Softw 2011;45:1–67.

34. Little RJA, Rubin DB. Statistical analysis with missing data, Second Edition. Jon Wiley & Sons, Inc ; 2002.

35. Khera R, Kondamudi N, Zhong L, Vaduganathan M, Parker J, Das SR, et al. Temporal Trends in Heart Failure Incidence Among Medicare Beneficiaries Across Risk Factor Strata, 2011 to 2016. JAMA Netw Open 2020;3:.

36. Tsao CW, Lyass A, Enserro D, Larson MG, Ho JE, Kizer JR, et al. Temporal Trends in the Incidence of and Mortality Associated With Heart Failure With Preserved and Reduced Ejection Fraction. JACC Heart Fail 2018;6:678–685.

37. Ford ES, Croft JB, Mannino DM, Wheaton AG, Zhang X, Giles WH. COPD surveillance--United States, 1999-2011. Chest 2013;144:284–305.

38. Maron BA, Choudhary G, Khan UA, Jankowich MD, McChesney H, Ferrazzani SJ, et al. Clinical profile and underdiagnosis of pulmonary hypertension in US veteran patients. Circ Heart Fail 2013;6:906–912.

39. Thiwanka Wijeratne D, Lajkosz K, Brogly SB, Diane Lougheed M, Jiang L, Housin A, et al. Increasing Incidence and Prevalence of World Health Organization Groups 1 to 4 Pulmonary Hypertension: A Population-Based Cohort Study in Ontario, Canada. Circ Cardiovasc Qual Outcomes 2018;11:.

40. Humbert M, Kovacs G, Hoeper MM, Badagliacca R, Berger RMF, Brida M, et al. 2022 ESC/ERS Guidelines for the diagnosis and treatment of pulmonary hypertension. Eur Heart J 2022;43:3618–3731.

41. D’Andrea A, Naeije R, Grünig E, Caso P, D’Alto M, Palma E Di, et al. Echocardiography of the pulmonary circulation and right ventricular function: exploring the physiologic spectrum in 1,480 normal subjects. Chest 2014;145:1071–1078.

42. Strange G, Playford D, Stewart S, Deague JA, Nelson H, Kent A, et al. Pulmonary hypertension: Prevalence and mortality in the Armadale echocardiography cohort. Heart 2012;98:.

43. Zierath R, Claggett B, Arthur V, Yang Y, Skali H, Matsushita K, et al. Changes in Pulmonary Artery Pressure Late in Life. J Am Coll Cardiol 2023;82:2179–2192.

44. Zhang L, Liu Y, Zhao S, Wang Z, Zhang M, Zhang S, et al. The Incidence and Prevalence of Pulmonary Hypertension in the COPD Population: A Systematic Review and Meta-Analysis. Int J Chron Obstruct Pulmon Dis 2022;17:1365–1379.

45. Cook DP, Xu M, Martucci VL, Annis JS, Aldrich MC, Hemnes AR, et al. Clinical insights into pulmonary hypertension in chronic obstructive pulmonary disease. Pulm Circ 2022;12:.

46. Lam CSP, Borlaug BA, Kane GC, Enders FT, Rodeheffer RJ, Redfield MM. Age-associated increases in pulmonary artery systolic pressure in the general population. Circulation 2009;119:2663–2670.

47. Fayngersh V, Drakopanagiotakis F, McCool FD, Klinger JR. Pulmonary hypertension in a stable community-based COPD population. Lung 2011;189:377–382.

48. D’Andrea A, Naeije R, Grünig E, Caso P, D’Alto M, Palma E Di, et al. Echocardiography of the pulmonary circulation and right ventricular function: exploring the physiologic spectrum in 1,480 normal subjects. Chest 2014;145:1071–1078.

49. Choudhary G, Jankowich M, Wu WC. Prevalence and clinical characteristics associated with pulmonary hypertension in African-Americans. PLoS One 2013;8:.

50. Yang BQ, Assad TR, O’Leary JM, Xu M, Halliday SJ, D’Amico RW, et al. Racial differences in patients referred for right heart catheterization and risk of pulmonary hypertension. Pulm Circ 2018;8:.

51. Brittain EL, Nwabuo C, Xu M, Gupta DK, Hemnes AR, Moreira HT, et al. Echocardiographic Pulmonary Artery Systolic Pressure in the Coronary Artery Risk Development in Young Adults (CARDIA) Study: Associations With Race and Metabolic Dysregulation. Journal of the American Heart Association: Cardiovascular and Cerebrovascular Disease 2017;6:.

52. Pugh ME, Robbins IM, Rice TW, West J, Newman JH, Hemnes AR. Unrecognized glucose intolerance is common in pulmonary arterial hypertension. J Heart Lung Transplant 2011;30:904– 911.

53. Robbins IM, Newman JH, Johnson RF, Hemnes AR, Fremont RD, Piana RN, et al. Association of the metabolic syndrome with pulmonary venous hypertension. Chest 2009;136:31–36.

54. Choudhary G, Jankowich M, Wu WC. Prevalence and clinical characteristics associated with pulmonary hypertension in African-Americans. PLoS One 2013;8:.

55. Huston JH, Maron BA, French J, Huang S, Thayer T, Farber-Eger EH, et al. Association of Mild Echocardiographic Pulmonary Hypertension With Mortality and Right Ventricular Function. JAMA Cardiol 2019;4:1112–1121.

56. Maron BA, Hess E, Maddox TM, Opotowsky AR, Tedford RJ, Lahm T, et al. Association of Borderline Pulmonary Hypertension With Mortality and Hospitalization in a Large Patient Cohort: Insights From the Veterans Affairs Clinical Assessment, Reporting, and Tracking Program. Circulation 2016;133:1240–1248.

57. D’Alto M, Bossone E, Opotowsky AR, Ghio S, Rudski LG, Naeije R. Strengths and weaknesses of echocardiography for the diagnosis of pulmonary hypertension. Int J Cardiol 2018;263:177–183.

58. Mukherjee M, Mercurio V, Balasubramanian A, Shah AA, Hsu S, Simpson CE, et al. Defining minimal detectable difference in echocardiographic measures of right ventricular function in systemic sclerosis. Arthritis Res Ther 2022;24:.

