## Supplementary material for "Incidence of Pulmonary Hypertension in the Echocardiography Referral Population": Online Supplement

**Table of Contents:**

Supplemental Methods: Pages 2 - 4

Tables E1-11: Pages 5 - 15

Figures E1-4: Pages 16 - 19

Supplemental References: Page 20

**Supplemental Methods**

*VA and Vanderbilt Cohorts*

We identified our exploratory cohort from participants in the VA Birth Cohort (1) an observational, prospective cohort consisting of 4.78 million U.S. VA patients born 1945-1965. Clinical and demographic data are extracted from the VA Corporate Data Warehouse and the VA electronic medical record Health Factor dataset. For the validation cohort we extracted data from Vanderbilt's Synthetic Derivative, a de-identified mirror image of the Vanderbilt electronic health record. For the VA cohort the start date of TTE report extraction (September 30, 1999) corresponds with the date on which electronic health records became available. For the Vanderbilt cohort, the start date (January 1, 1994) corresponds with the date when TTE reports became digitized.

*Exposures, Covariates, and Echocardiographic Data*

HF and COPD are the most common causes of PH in the Unites States and among patients seeking care at tertiary care centers, therefore these were selected as our primary exposures of interest (2). The diagnostic codes utilized to determine the presence of COPD and HF are detailed in **Table E11**. A patient was considered to have COPD or HF if they had two outpatient ICD codes or one inpatient ICD code.  Smoking data has not been well validated across the study period in the Vanderbilt cohort and therefore was not included in predictive models for the validation cohort. FIB-4 data was not available in the Vanderbilt dataset. Cocaine abuse was included as a covariate due to evidence of its association with elevated PASP (3). Hypertension was defined by the presence of a prescription for an anti-hypertensive medication, which was further categorized as controlled or uncontrolled based on the average of the 3 reported blood pressure measures closest to the date of TTE. Controlled hypertension was defined by an average blood pressure of <140/90 while uncontrolled hypertension was defined by an average blood pressure of ≥140/90 mmHg. We considered inclusion of hyperthyroidism and supplemental oxygen use, however, ultimately did not add in these variables due to low prevalence (<1.5%) in both cohorts.

We used validated natural language processing tools to extract echocardiographic measurements from VA and Vanderbilt TTE reports (4, 5) We calculated missing PASP measurements by the simplified Bernoulli equation, using tricuspid regurgitant velocity and estimated right atrial pressure (RAP). We assumed a RAP of 5 mmHg for missing values as this was median value in both cohorts and a conservative assumption consistent with previous population-based studies (6, 7). We excluded PASP estimates outside a physiological range (<8 mm Hg or >159 mm Hg) based on clinical experience. Heart rate at the time of TTE could not be abstracted with the current natural language processing tool.

*Outcome Analysis*

Patients with only one measured PASP value were assumed to never develop PH in the primary analysis. This assumption was made to mirror clinical practice and to derive a conservative estimate of PH incidence in this cohort. We also completed a subsequent sensitivity analysis conducted only among individuals with a second TTE to describe the range of potential IRs if all patients were to have follow up. To compare between TTE-based and ICD-based IRs, we calculated the PH IR using ICD-9 and ICD-10 diagnostic codes (416.0, 416.1, 417.8,

416.9, I27.0, I27.1, I27.2, I27.8, I27.9). One inpatient diagnostic code was required to establish the diagnosis of PH. This was the same definition used in the prior study by Wijeratne et al (8).

Mortality data in the VA cohort was determined from the VA vital status file, which is compiled from multiple VA and non-VA data sources in the VBA Integrated Benefits System (IBS) Death File (formally known as the BIRLS Death file), Medicare Vital Status File, and the Social Security Administration (SSA) Death Master File 16. Mortality data in the Vanderbilt cohort was determined from the Social Security Death Index (SSDI). The Vanderbilt mortality analysis truncated at the end of 2017, after which the synthetic derivative is no longer linked to the SSDI.

| Table E1: Characteristics of Patients with and without a PASP repeat estimate in the VA Cohort | | | |
|---|---|---|---|
| | **Overall** (n=24506) | **One PASP** (n=158060) | **>1 PASP** (n=87007) |
| **Incident PH** | 38882 (15.9%) | 0 (0%) | 38882 (44.7%) |
| **Age, years (median [IQR])** | 59.9 [55.1, 64.1] | 60.1 [55.0, 64.2] | 59.8 [55.2, 63.8] |
| **Male Sex (%)** | 229637 (93.7) | 147300 (93.2) | 82337 (94.6) |
| **BMI (median [IQR])** | 29.5 [25.9, 33.9] | 29.4 [25.8, 33.7] | 29.8 [26.2, 34.2] |
| **Race/Ethnicity (%)** | | | |
| Other | 7986 (3.3) | 5146 (3.3) | 2840 (3.3) |
| Black | 48396 (19.7) | 31180 (19.7) | 17216 (19.8) |
| Hispanic | 12770 (5.2) | 8237 (5.2) | 4533 (5.2) |
| White | 165165 (67.4) | 105580 (66.8) | 59585 (68.5) |
| **HF (%)** | 44637 (18.2) | 22457 (14.2) | 22180 (25.5) |
| **COPD (%)** | 68877 (28.1) | 43197 (27.3) | 25680 (29.5) |
| **Alcohol abuse (%)** | 62668 (25.6) | 40863 (25.9) | 21805 (25.1) |
| **Cocaine abuse (%)** | 19880 (8.1) | 12730 (8.1) | 7150 (8.2) |
| **Scleroderma (%)** | 6942 (2.8) | 4134 (2.6) | 2808 (3.2) |
| **Smoking (%)** | | | |
| Current Smoker | 77130 (31.5) | 50572 (32.0) | 26558 (30.5) |
| Former Smoker | 57897 (23.6) | 36244 (22.9) | 21653 (24.9) |
| Never Smoker | 56503 (23.1) | 37156 (23.5) | 19347 (22.2) |
| **Hypertension (%)** | | | |
| None | 7757 (3.2) | 6606 (4.2) | 1151 (1.3) |
| Controlled | 183639 (74.9) | 116790 (73.9) | 66849 (76.8) |
| Uncontrolled | 45609 (18.6) | 28909 (18.3) | 16700 (19.2) |
| **Diabetes Mellitus (%)** | 100935 (41.2) | 60372 (38.2) | 40563 (46.6) |
| **Hepatitis C Infection (%)** | 28558 (11.7) | 18132 (11.5) | 10426 (12.0) |
| **FIB-4 >3.25** | 13980 (5.7) | 9262 (5.9) | 4718 (5.4) |
| **eGFR (median [IQR])** | 85.0 [71.0, 101.0] | 86.0 [73.0, 103.0] | 85.0 [69.0, 99.0] |
| **Hgb (median [IQR])** | 14.2 [12.9, 15.2] | 14.2 [12.9, 15.2] | 14.1 [12.8, 15.2] |

BMI = Body Mass Index (in $kg/m^2$)
HF = Heart Failure
eGFR = estimated Glomerular Filtration Rate (in $mL/min/1.73m^2$)
Hgb = Hemoglobin (in g/dL)

| Table E2: Characteristics of Patients by Time between Echocardiograms | | | | | |
|---|---|---|---|---|---|
| | **>1 PASP** (n=87007) | **<2 years** (n=35628) | **2 – 4 years** (n=22573) | **4 – 6 years** (n=12617) | **>6 years** (n=16189) |
| **Incident PH** | 38882 (44.7%) | 16975 (47.6) | 9687 (42.9) | 5376 (42.6) | 6844 (42.3) |
| **Age, years (median [IQR])** | 59.8 [55.2, 63.8] | 60.5 [56.1, 64.5] | 60.5 [56.1, 64.6] | 59.9 [55.2, 63.7] | 57.0 [52.7, 60.8] |
| **Male Sex (%)** | 82337 (94.6) | 33935 (95.2) | 21362 (94.6) | 11928 (94.5) | 15112 (93.3) |
| **BMI (median [IQR])** | 29.8 [26.2, 34.2] | 29.5 [25.8, 33.9] | 30.0 [26.4, 34.3] | 29.9 [26.4, 34.3] | 30.0 [26.5, 34.2] |
| **Race/Ethnicity (%)** | | | | | |
| Other | 2840 (3.3) | 1112 (3.1) | 726 (3.2) | 399 (3.2) | 603 (3.7) |
| Black | 17216 (19.8) | 6779 (19.0) | 4462 (19.8) | 2525 (20.0) | 3450 (21.3) |
| Hispanic | 4533 (5.2) | 1767 (5.0) | 1130 (5.0) | 724 (5.7) | 912 (5.6) |
| White | 59585 (68.5) | 24648 (69.2) | 15562 (68.9) | 8602 (68.2) | 10773 (66.5) |
| **HF (%)** | 22180 (25.5) | 12109 (34.0) | 5296 (23.5) | 2428 (19.2) | 2347 (14.5) |
| **COPD (%)** | 25680 (29.5) | 11830 (33.2) | 6749 (29.9) | 3455 (27.4) | 3646 (22.5) |
| **Alcohol abuse (%)** | 21805 (25.1) | 9701 (27.2) | 5570 (24.7) | 3021 (23.9) | 3513 (21.7) |
| **Cocaine abuse (%)** | 7150 (8.2) | 3131 (8.8) | 1744 (7.7) | 987 (7.8) | 1288 (8.0) |
| **Scleroderma (%)** | 2808 (3.2) | 1217 (3.4) | 744 (3.3) | 405 (3.2) | 442 (2.7) |
| **Smoking (%)** | | | | | |
| Current Smoker | 26558 (30.5) | 10802 (30.3) | 6782 (30.0) | 3785 (30.0) | 5189 (32.1) |
| Former Smoker | 21653 (24.9) | 9035 (25.4) | 5740 (25.4) | 3145 (24.9) | 3733 (23.1) |
| Never Smoker | 19347 (22.2) | 7632 (21.4) | 5070 (22.5) | 2944 (23.3) | 3701 (22.9) |
| **Hypertension (%)** | | | | | |
| None | 1151 (1.3) | 453 (1.3) | 319 (1.4) | 187 (1.5) | 192 (1.2) |
| Controlled | 66849 (76.8) | 27827 (78.1) | 17308 (76.7) | 9634 (76.4) | 12080 (74.6) |
| Uncontrolled | 16700 (19.2) | 6514 (18.3) | 4463 (19.8) | 2478 (19.6) | 3245 (20.0) |
| **Diabetes Mellitus (%)** | 40563 (46.6) | 16489 (46.3) | 10355 (45.9) | 5967 (47.3) | 7752 (47.9) |
| **Hepatitis C Infection (%)** | 10426 (12.0) | 4667 (13.1) | 2558 (11.3) | 1380 (10.9) | 1821 (11.2) |
| **FIB-4 >3.25** | 4718 (5.4) | 2705 (7.6) | 1070 (4.7) | 504 (4.0) | 439 (2.7) |
| **eGFR (median [IQR])** | 85.0 [69.0, 99.0] | 84 [68, 99] | 85 [70, 99] | 85 [71, 101] | 86 [72, 99] |
| **Hgb (median [IQR])** | 14.1 [12.8, 15.2] | 13.9 [12.4, 15.0] | 14.2 [12.9, 15.2] | 14.2 [13.1, 15.2] | 14.4 [13.3, 15.3] |

BMI = Body Mass Index (in $kg/m^2$)
HF = Heart Failure
eGFR = estimated Glomerular Filtration Rate (in $mL/min/1.73m^2$)
Hgb = Hemoglobin (in g/dL)

| Table E3: Pulmonary Hypertension IRs in the VA and Vanderbilt Cohorts restricted to patients with a second PASP measurement, stratified by HF/COPD Status and within HF by LVEF | | | | | | |
|---|---|---|---|---|---|---|
| | **VA PH IRs** | | | **Vanderbilt PH IRs** | | |
| | **N** | **Incident PH** | **Rate/1000 PY** | **N** | **Incident PH** | **Rate/1000 PY** |
| **Total** | 87007 | 38882 | 65.9 (65.2, 66.5) | 21653 | 8061 | 69.1 (67.6, 70.6) |
| **HF-/COPD-** | 48874 | 19111 | 51.0 (50.2, 51.7) | 13328 | 4199 | 51.3 (49.8, 52.9) |
| **HF-/COPD+** | 15953 | 7128 | 69.9 (68.3, 71.5) | 1329 | 555 | 93.7 (86.1, 101.7) |
| **HF+/COPD-** | 12453 | 6856 | 98.2 (95.9, 100.6) | 5542 | 2524 | 104.3 (100.3, 108.5) |
| **HF+/COPD+** | 9727 | 5787 | 133.4 (130.0, 136.9) | 1454 | 783 | 168.3 (156.8, 180.4) |
| **HF+/LVEF<50** | 13791 | 7966 | 115.5 (113.0, 118.1) | 3006 | 1478 | 128.4 (122.0, 135.1) |
| **HF+/LVEF≥50%** | 8389 | 4677 | 105.8 (102.8, 108.8) | 3843 | 1751 | 105.2 (100.4, 110.2) |

HF = Heart Failure
COPD = Chronic Obstructive Pulmonary Disease
LVEF = Left Ventricular Ejection Fraction

| Table E4: Pulmonary Hypertension Incident Rates by Year of Baseline Echocardiogram in the VA cohort | | | |
|---|---|---|---|
| **Years** | **Overall Population** | **# of Incident PH** | **Rate/1000 PY** |
| 2000-2005 | 32866 | 1651 | 26.8 (25.5, 28.1) |
| 2005-2010 | 91543 | 5622 | 26.4 (25.7, 27.1) |
| 2010-2015 | 125551 | 7799 | 23.9 (23.4, 24.4) |
| >2015 | 47736 | 5123 | 25.9 (25.2, 26.6) |

| Table E5: Pulmonary Hypertension Incidence Rates in the VA and VUMC Cohorts with PH defined by echocardiographic PASP greater than 40 mmHg | | | | | | |
|---|---|---|---|---|---|---|
| | VA PH Incidence Rates | | | VUMC PH Incidence Rates | | |
| | **N** | **Incident PH** | **Rate/1000 PY** | **N** | **Incident PH** | **Rate/1000 PY** |
| **Total** | 245067 | 25710 | 14.0 (13.9, 14.2) | 77548 | 5043 | 15.3 (14.9, 15.8) |
| **HF-/COPD-** | 152456 | 11752 | 9.4 (9.2, 9.5) | 55258 | 2418 | 9.6 (9.2, 10.0) |
| **HF-/COPD+** | 47974 | 4616 | 14.1 (13.7, 14.5) | 5687 | 355 | 19.3 (17.4, 21.4) |
| **HF+/COPD-** | 23734 | 5022 | 33.9 (33.0, 34.8) | 12741 | 1710 | 35.5 (33.8, 37.2) |
| **HF+/COPD+** | 20903 | 4320 | 42.1 (40.8, 43.3) | 3862 | 560 | 52.7 (48.4, 57.2 |
| **HF+/LVEF<50** | 21655 | 5234 | 44.2 (43.0, 45.4) | 5129 | 893 | 53.1 (49.7, 56.7) |
| **HF+/LVEF>=50** | 22982 | 4108 | 31.0 (30.1, 32.0) | 11090 | 1319 | 32.5 (30.8, 34.3) |

HF = Heart Failure
COPD = Chronic Obstructive Pulmonary Disease
LVEF = Left Ventricular Ejection Fraction

| Table E6: Incident PH Diagnosis by TTE compared to ICD-Coding in the VA Cohort and Vanderbilt Cohort | | | | | | | | | |
|---|---|---|---|---|---|---|---|---|---|
| **VA Cohort** | | **Incident PH by ICD-Code*** | | | **Vanderbilt Cohort** | | **Incident PH by ICD-Code*** | | |
| | | No | Yes | Sum | | | No | Yes | Sum |
| **Incident PH by TTE** | No | 214392 | 4516 | 218908 | **Incident PH by TTE** | No | 67790 | 1697 | 69487 |
| | Yes | 21415 | 4744 (18%) | 26159 | | Yes | 6494 | 1567 (19%) | 8061 |
| | Sum | 235807 | 9260 | | | Sum | 74284 | 3264 | 77548 |
| *ICD codes included: 416.0, 416.1, 416.8, 416.9, I27.0, I27.1, I27.2, I27.8, I27.9 | | | | | | | | | |

| Table E7: Pulmonary Hypertension Incidence Rates in the VA and Vanderbilt Cohorts (PH defined by ICD-coding) | | | | | | |
|---|---|---|---|---|---|---|
| | VA PH Incidence Rates† | | | Vanderbilt PH Incidence Rates | | |
| | **N** | **Incident PH** | **Rate/1000 PY** | **N** | **Incident PH** | **Rate/1000 PY** |
| **Total** | 245058 | 9260 | 7.5 (7.3, 7.6) | 76277 | 1993 | 5.9 (5.7, 8.5) |
| **HF-/COPD-** | 152448 | 3282 | 3.9 (3.8, 4.0) | 55039 | 722 | 2.8 (2.6, 3.0) |
| **HF-/COPD+** | 47973 | 1859 | 8.6 (8.2, 9.0) | 5557 | 157 | 8.4 (7.2, 9.8) |
| **HF+/COPD-** | 23734 | 1796 | 16.5 (15.8, 17.3) | 12091 | 789 | 16.1 (15.0, 17.3) |
| **HF+/COPD+** | 20903 | 2323 | 31.9 (30.6, 33.2) | 3550 | 325 | 31.3 (28.0, 34.8) |

*ICD codes included: 4160, 4161, 4168, 4169, I270, I271, I272, I278, I279
†Truncated at 12/2016
Patients with code-based PH prior to study entry were excluded

| Table E8: Predictors of Incident PH in the VA and Vanderbilt Cohorts (Sensitivity Analysis of PASP greater than or equal to 40 mmHg) | | | | |
|---|---|---|---|---|
| **Characteristic** | **VA Cohort** | | **Vanderbilt Cohort** | |
| | **Hazard Ratio [95% CI]** | | **Hazard Ratio [95% CI]** | |
| | **Baseline** | **Time updated *** | **Baseline** | **Time updated *** |
| **HF-/COPD-** | 1 | 1 | 1 | 1 |
| **HF-/COPD+** | 1.36 (1.31-1.41) | 1.62 (1.55-1.69) | 1.53 (1.36-1.71) | 1.85 (1.64-2.08) |
| **HF+/COPD-** | 2.67 (2.58-2.76) | 4.01 (3.85-4.17) | 2.48 (2.33-2.65) | 3.52 (3.28-3.77) |
| **HF+/COPD+** | 2.94 (2.83-3.05) | 5.32 (5.12-5.53) | 3.01 (2.73-3.32) | 4.82 (4.40-5.28) |
| **Age, years (per SD, 6.4)** | 1.13 (1.12-1.15) | 1.07 (1.06-1.09) | 1.41 (1.26-1.36) | 1.16 (1.12-1.20) |
| **Male Sex** | 1.52 (1.42-1.62) | 1.40 (1.30-1.50) | 1.14 (1.08-1.21) | 1.11 (1.04-1.18) |
| **BMI, kg/m$^2$ (per SD, 6.4)** | 1.01 (0.99-1.02) | 0.97 (0.96-0.99) | 1.00 (0.97-1.03) | 1.00 (0.97-1.03) |
| **Race/Ethnicity (vs. Non-Hispanic White)** | | | | |
| Other | 0.98 (0.91-1.05) | 1.01 (0.94-1.09) | 0.78 (0.66-0.93) | 0.79 (0.65-0.95) |
| Non-Hispanic Black | 1.11 (1.07-1.15) | 1.12 (1.09-1.17) | 1.35 (1.24-1.46) | 1.32 (1.21-1.44) |
| Hispanic | 0.93 (0.88-0.99) | 0.96 (0.90-1.03) | 0.73 (0.54-0.99) | 0.76 (0.55-1.03) |
| **Hypertension (vs. No Hypertension)** | | | | |
| *Controlled Hypertension* | 2.25 (1.91-2.65) | 1.84 (1.56-2.17) | 1.54 (1.28-1.87) | 1.31 (1.08-1.59) |
| *Uncontrolled Hypertension* | 2.76 (2.35-3.25) | 2.10 (1.77-2.48) | 1.65 (1.36-2.00) | 1.35 (1.11-1.65) |
| **Diabetes Mellitus** | 1.40 (1.36-1.43) | 1.21 (1.18-1.25) | 1.37 (1.28-1.47) | 1.13 (1.05-1.21) |
| **Veterans with HIV** | 1.12 (1.00-1.24) | 1.03 (0.92-1.16) | 0.87 (0.69-1.08) | 0.83 (0.66-1.04) |
| **Hepatitis C Infection** | 1.24 (1.20-1.29) | 1.22 (1.17-1.27) | 1.48 (1.26-1.73) | 1.45 (1.23-1.70) |
| **Smoking Status (vs. Never)** | | | - | - |
| *Current* | 1.22 (1.18-1.27) | 1.12 (1.08-1.16) | - | - |
| *Former* | 1.09 (1.05-1.13) | 1.04 (1.00-1.08) | - | - |
| **Liver Fibrosis (FIB-4 >3.25 vs. <u><</u>325)** | 1.33 (1.26-1.41) | 1.37 (1.29-1.46) | - | - |
| **eGFR, mL/min/1.73m$^2$ (per SD, 2.9)** | 0.87 (0.86-0.88) | 0.69 (0.68-0.70) | 0.78 (0.75-0.82) | 0.59 (0.56-0.62) |
| **Alcohol abuse** | 1.05 (1.01-1.08) | 1.04 (1.00-1.07) | 1.00 (0.86-1.17) | 0.99 (0.85-1.16) |
| **Scleroderma** | 1.16 (1.08-1.24) | 1.15 (1.07-1.24) | 1.33 (1.18-1.50) | 1.29 (1.14-1.45) |
| **Cocaine abuse** | 1.01 (0.96-1.06) | 0.98 (0.93-1.03) | 1.24 (0.96-1.60) | 1.18 (0.91-1.54) |
| **Atrial fibrillation** | 1.65 (1.60-1.70) | 1.49 (1.44-1.54) | 1.65 (1.55-1.76) | 1.52 (1.42-1.62) |
| **Hgb, mg/dL (per SD, 1.9)** | 0.82 (0.81-0.83) | 0.88 (0.87-0.89) | 0.81 (0.79-0.84) | 0.85 (0.83-0.88) |

*HF, COPD, and eGFR and are time updated. Other variables measured solely at baseline
HF = Heart Failure
COPD = Chronic Obstructive Pulmonary Disease
BMI = Body Mass Index
eGFR = estimated Glomerular Filtration Rate
Hgb = Hemoglobin

| Table E9: Mortality rates after incident PH by HF and COPD status in the VA and Vanderbilt Cohorts | | | | | | |
|---|---|---|---|---|---|---|
| | VA Cohort | | | Vanderbilt Cohort* | | |
| | **# of Incident PH** | **Mortality Events** | **Rate/1000 PY** | **# of Incident PH** | **Mortality Events** | **Rate/1000 PY** |
| **HF-/COPD-** | 18517 | 5872 | 67.7 (66.0, 69.5) | 1861 | 198 | 27.9 (24.2, 32.0) |
| **HF-/COPD+** | 7122 | 3039 | 107.9 (104.1, 111.9) | 386 | 64 | 55.9 (43.3, 70.8) |
| **HF+/COPD-** | 7149 | 3616 | 114.1 (110.5, 117.9) | 2455 | 435 | 50.8 (46.2, 55.8) |
| **HF+/COPD+** | 6005 | 3711 | 172.4 (167.0, 178.0) | 1156 | 248 | 80.5 (70.9, 90.9) |
| **Total** | 38793 | 16238 | 96.6 (95.1, 98.1) | 5858 | 945 | 47.6 (44.6, 50.7) |

*Vanderbilt cohort truncated at the end of 2017
HF = Heart Failure
COPD = Chronic Obstructive Pulmonary Disease

| Table E10: Mortality rates by PASP Category in the VA and Vanderbilt Cohorts | | | | | | |
|---|---|---|---|---|---|---|
| | VA Cohort | | | Vanderbilt Cohort* | | |
| | # of Incident PH | Mortality Events | Rate/1000 PY | # of Incident PH | Mortality Events | Rate/1000 PY |
| **No PH** | 206108 | 58152 | 42.5 (42.2, 42.9) | 55000 | 2796 | 16.4 (15.8, 17.0) |
| **[35, 45)** | 26188 | 9684 | 79.5 (78.0, 81.1) | 4168 | 608 | 40.0 (37.4, 43.8) |
| **[45, 55)** | 7789 | 3965 | 133.2 (129.1, 137.4) | 1114 | 224 | 66.2 (57.9, 75.2) |
| **[55, 65)** | 2827 | 1579 | 168.1 (159.9, 176.5) | 398 | 76 | 74.9 (59.3, 93.1) |
| **65+** | 1989 | 1010 | 141.2 (132.6, 150.1) | 178 | 37 | 78.3 (55.7, 106.3) |
| *Vanderbilt cohort truncated at the end of 2017 | | | | | | |

| Table E11: Diagnostic Codes for Determination of HF and COPD status | | |
|---|---|---|
| | ICD-9 codes | ICD-10 codes |
| Heart Failure | 40201, 40211, 40291, 40401, 40403, 40411, 40413, 40491, 40493, 4280, 4281, 42821, 42822, 42823, 42831, 42832, 42833, 42841, 42842, 42843, 4289, 42820, 42830, 42840 | I110, I130, I132, I501, I5020, I5021, I5022, I5023, I5030, I5031, I5032, I5033, I5040, I5041, I5042, I5043, I50810, I50811, I50812, I50813, I50814, I5082, I5083, I5084, I5089, I509 |
| COPD^ | 490, 491.0, 491.1, 491.2, 491.20, 491.21, 491.22, 491.8, 491.9, 496, 492.8, 492.0 | J40.*, J41.*, J42.*, J43.*, J44.* |
| *Indicating all numbers after decimal (0-9)<br>NOTE: Diagnosis established by the presence of one inpatient ICD 9/10 code or two outpatient ICD 9/10 codes, at the time of the second outpatient ICD code. | | |

**Figure E1:** Time to Development of Incident PH

Displayed are the Kaplan-Meier Curves for time to development of incident PH in the VA Cohort (A) and the Vanderbilt Cohort (B), with stratification by HF and COPD status. Concurrent HF and COPD are associated with the highest unadjusted rate of incident PH, followed by HF, then COPD, with the lowest rate noted in patients without HF or COPD. Results are similar in both cohorts.

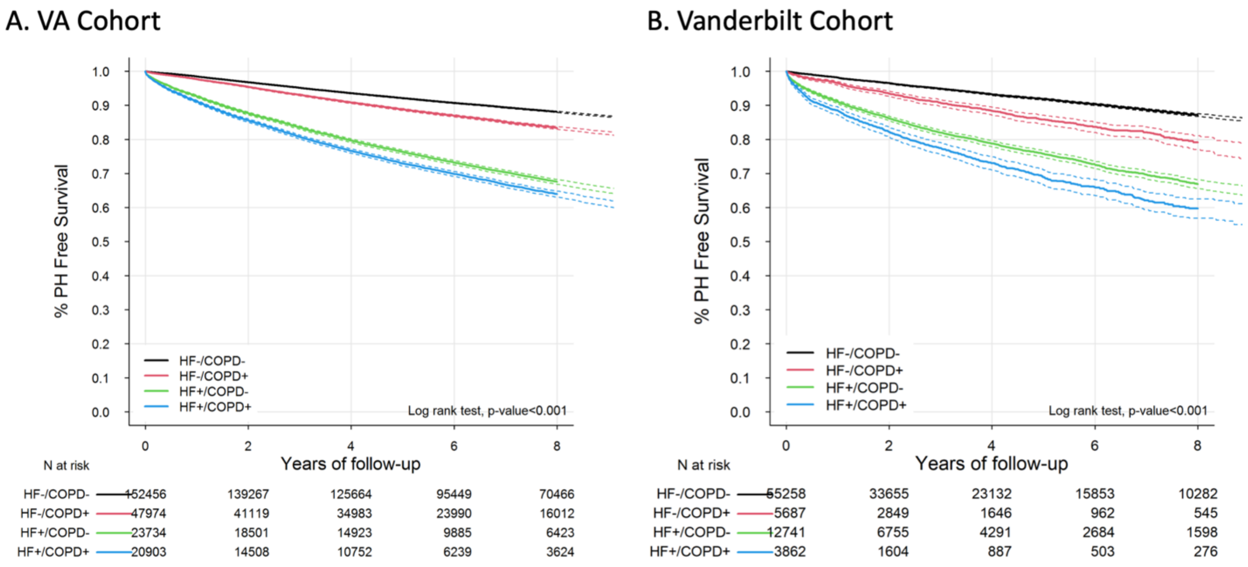

**Figure E2:** Survival following Incident PH by PASP Category

Displayed are Kaplan-Meier time-to-event curves for survival in the VA cohort, with stratification by PASP categories (8-35; 35-45; 45-55; 55-65; and above 65). We assessed the differential effect of PASP on survival in the full cohort (A), patients with HF and no COPD (B), patients with COPD and no HF (C), and patients with neither COPD nor HF (D). We found similar effects in all cohorts examined, with the rate of mortality increasing as PASP category increased until the 45-55 category, with a similar rate of mortality among the 45-55, 55-65, and above 65 groups.

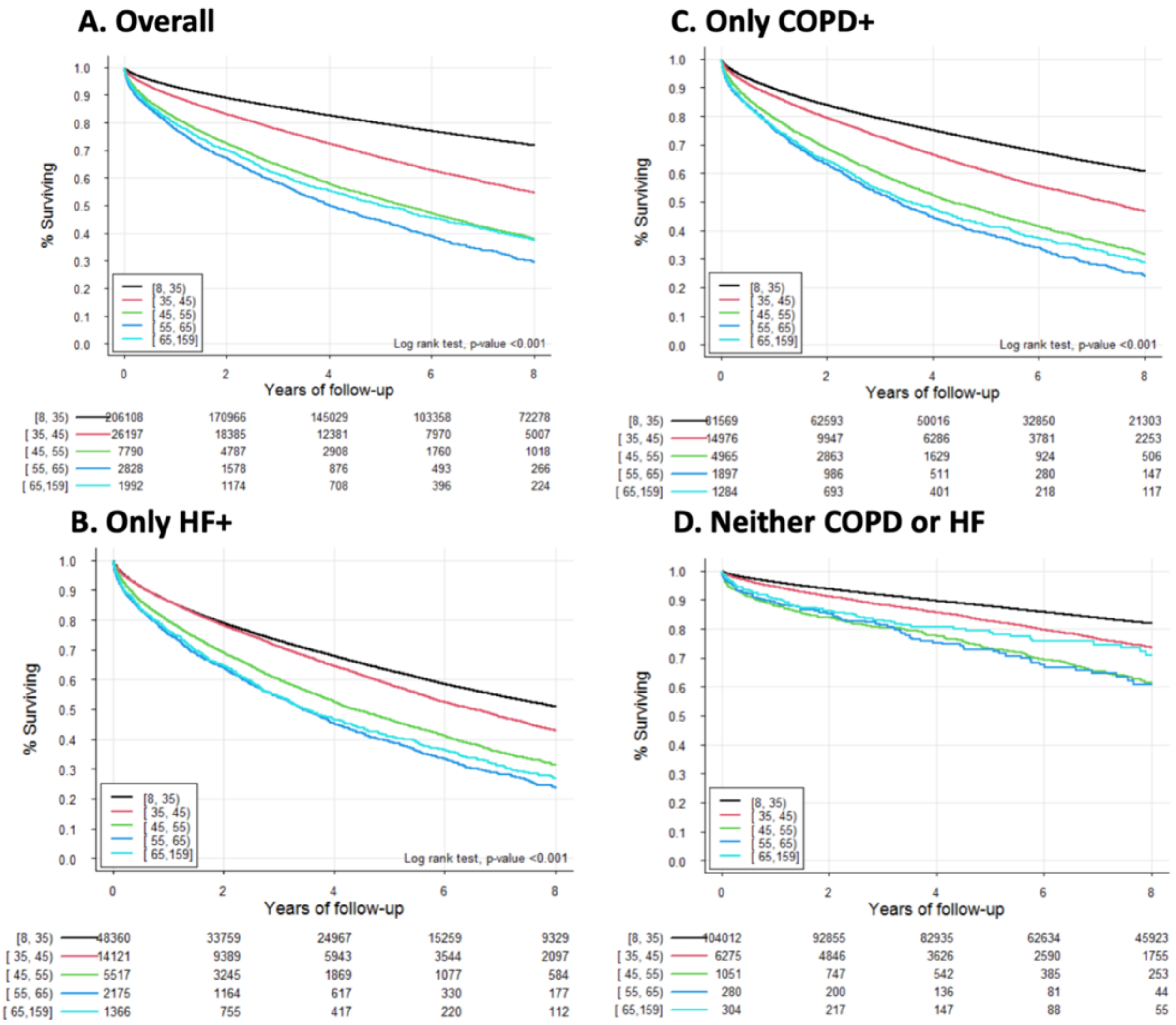

**Figure E3:** Survival following Incident PH by time between Echocardiograms

Displayed are Kaplan-Meier time-to-event curves for survival in the VA cohort, with stratification by PASP categories (35-45; 45-55; 55-65; and above 65) in patients with two or fewer years between TTEs (A), four or fewer years between TTEs (B), six or fewer years between TTEs (C) and eight or fewer years between TTEs (D). We noted similar rates of mortality by PASP category regardless of time between echocardiograms.

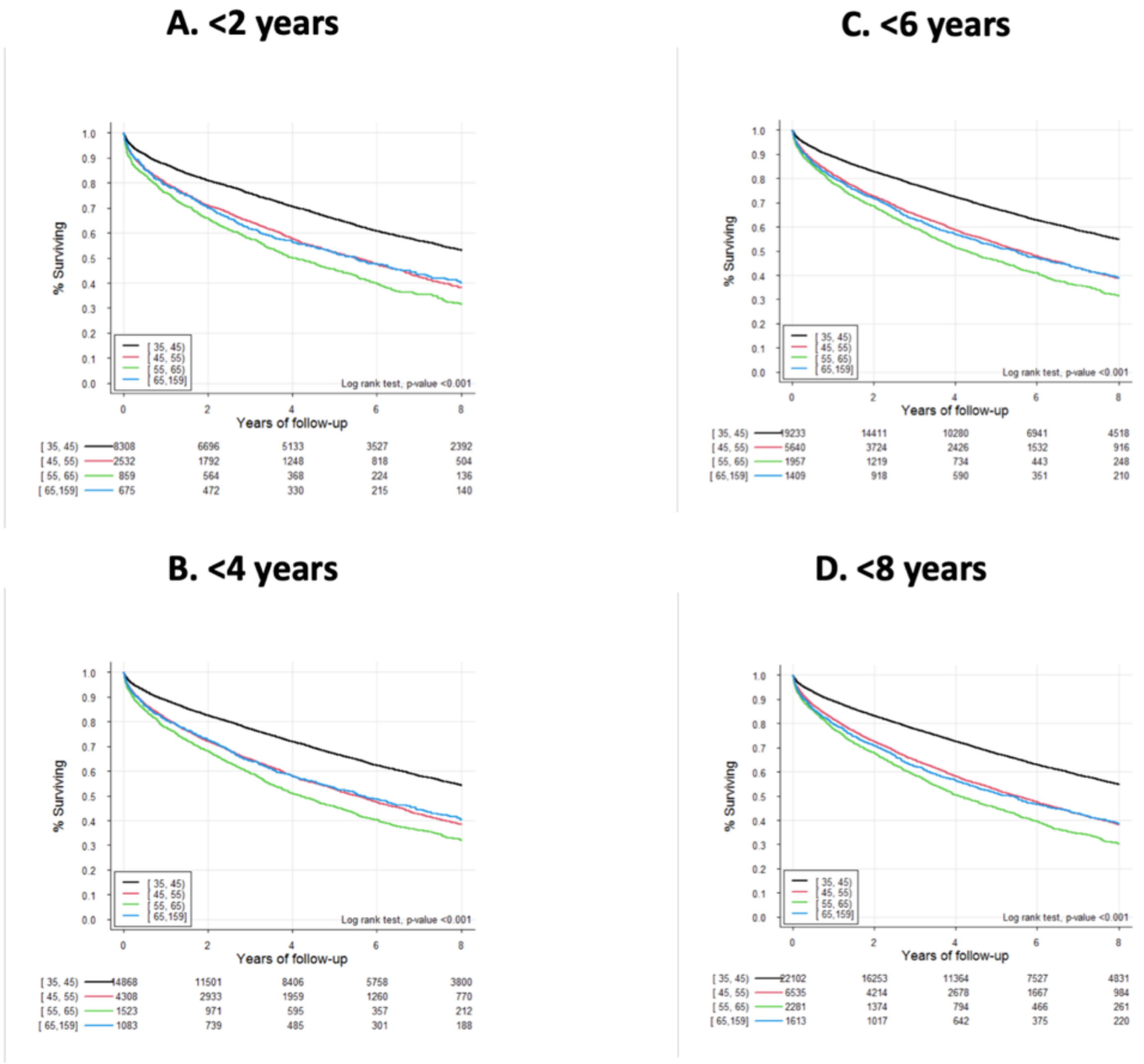

**Figure E4:** Mortality hazard by PASP at incident PH diagnosis (Vanderbilt Cohort)

We evaluated the relationship between PASP at the time of echocardiographic incident PH diagnosis and hazard of mortality by restricted cubic splines with up to 3 knots in the Vanderbilt cohort. We found that the relative hazard of mortality increases linearly as PASP increases from 35 mmHg up to approximately 45mmHg, above which hazard of mortality continues to increase at a substantially slower rate (A). We then stratified these results by gender and found that the hazard of mortality appears to continue increasing once PASP increases above 45 for women, but plateaus and does not further increase for men. These results are similar to the findings in the VA cohort (Figure 2).

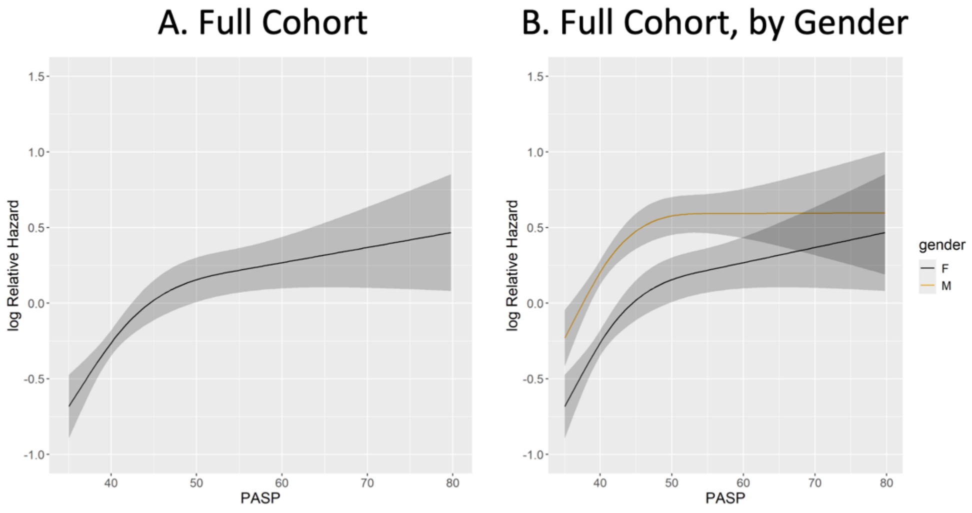

**Online Supplement References**


1. Sarkar S, Esserman DA, Skanderson M, Levin FL, Justice AC, Lim JK. Disparities in hepatitis C testing in U.S. veterans born 1945-1965. *J Hepatol* 2016;65:259–265.
2. Hoeper MM, Humbert M, Souza R, Idrees M, Kawut SM, Sliwa-Hahnle K, *et al.* A global view of pulmonary hypertension. *Lancet Respir Med* 2016;
3. Alzghoul BN, Abualsuod A, Alqam B, Innabi A, Palagiri DR, Gheith Z, *et al.* Cocaine Use and Pulmonary Hypertension. *American Journal of Cardiology* 2020;125:.
4. Patterson O V., Freiberg MS, Skanderson M, Fodeh JS, Brandt CA, DuVall SL. Unlocking echocardiogram measurements for heart disease research through natural language processing. *BMC Cardiovasc Disord* 2017;17:.
5. Wells QS, Farber-Eger E, Crawford DC. Extraction of echocardiographic data from the electronic medical record is a rapid and efficient method for study of cardiac structure and function. *J Clin Bioinforma* 2014;4:.
6. Lam CSP, Roger VL, Rodeheffer RJ, Borlaug BA, Enders FT, Redfield MM. Pulmonary hypertension in heart failure with preserved ejection fraction: a community-based study. *J Am Coll Cardiol* 2009;53:1119–1126.
7. Choudhary G, Jankowich M, Wu WC. Elevated pulmonary artery systolic pressure predicts heart failure admissions in African Americans jackson heart study. *Circ Heart Fail* 2014;7:558–564.
8. Thiwanka Wijeratne D, Lajkosz K, Brogly SB, Diane Lougheed M, Jiang L, Housin A, *et al.* Increasing Incidence and Prevalence of World Health Organization Groups 1 to 4 Pulmonary Hypertension: A Population-Based Cohort Study in Ontario, Canada. *Circ Cardiovasc Qual Outcomes* 2018;11:.
